# Development and Validation of a Pan-Cancer Stromal Activity Score for Predicting Prognosis and Immunotherapy Response

**DOI:** 10.64898/2026.07.21.26358625

**Authors:** Keran Sun, Keqi Jia

## Abstract

**Background:** The tumor microenvironment (TME) plays a critical role in cancer progression and treatment response. Stromal components, including cancer-associated fibroblasts (CAFs), extracellular matrix (ECM), and angiogenesis, contribute to tumor aggressiveness. However, a comprehensive stromal activity score integrating multiple stromal dimensions for pan-cancer prognosis prediction is lacking.

**Methods:** We developed a Stromal Activity Score (SAS) integrating five stromal dimensions: CAF signature (12 genes), ECM remodeling (15 genes), TGF-β signaling (13 genes), angiogenesis (12 genes), and complement activation (11 genes). SAS was calculated using single-sample Gene Set Enrichment Analysis (ssGSEA) on TCGA pan-cancer data comprising 1,303 samples across 12 cancer types. Prognostic value was evaluated using Kaplan-Meier analysis and Cox regression. Immunotherapy response prediction was validated in two independent cohorts (IMvigor210, n=88; Liu2019, n=105).

**Results:** Pan-cancer Cox regression demonstrated a significant association between SAS and overall survival (HR = 1.165, 95% CI: 1.065–1.275, P = 0.001). Per-cancer analysis identified BRCA (HR = 1.942, P = 0.022), STAD (HR = 1.684, P = 0.024), and LUSC (HR = 1.552, P = 0.038) as significant, though none survived FDR correction. SAS correlated strongly with ESTIMATE Stromal Score (Spearman ρ = 0.835) and moderately with Immune Score (ρ = 0.396). Immunotherapy validation showed consistent trends (IMvigor210: AUC = 0.602; Liu2019: AUC = 0.617). Time-dependent ROC analysis showed 1-year AUC = 0.596, 3-year = 0.579, 5-year = 0.559. Leave-one-out analysis identified angiogenesis removal as enhancing prognostic signal (HR = 3.737, P = 0.0002). Three distinct TME subtypes were identified with differential SAS profiles.

**Conclusions:** SAS is a novel pan-cancer stromal activity score that captures TME biology with strong construct validity. Its clinical utility as a standalone biomarker remains modest, but it may complement existing immunotherapy biomarkers.

## INTRODUCTION

The tumor microenvironment (TME) is a complex ecosystem comprising cancer cells, immune cells, stromal cells, and extracellular matrix components [1]. Stromal components, particularly cancer-associated fibroblasts (CAFs), play crucial roles in tumor progression, metastasis, and treatment resistance [2]. Recent single-cell RNA sequencing (scRNA-seq) studies have revealed extensive heterogeneity within stromal populations, with distinct CAF subtypes exhibiting different functional properties [3].

The classical CAF classification includes myofibroblastic CAFs (myCAF), inflammatory CAFs (iCAF), and antigen-presenting CAFs (apCAF) [4]. Each subtype contributes differently to tumor biology: myCAFs promote ECM remodeling and stiffness, iCAFs create an immunosuppressive niche through cytokine secretion, and apCAFs modulate immune responses through antigen presentation [5].

Beyond CAFs, other stromal components including ECM remodeling, angiogenesis, and complement activation also contribute to tumor progression [6,7]. ECM remodeling creates a physical barrier that impedes immune cell infiltration [8]. Angiogenesis promotes tumor growth and metastasis [9]. Complement activation can either promote or suppress antitumor immunity depending on context [10–13].

However, most studies focus on individual stromal components rather than integrating multiple stromal dimensions. A comprehensive stromal activity score could provide better insights into TME biology and predict treatment response more accurately.

In this study, we developed a pan-cancer Stromal Activity Score (SAS) integrating five stromal dimensions: CAF signature, ECM remodeling, TGF-β signaling, angiogenesis, and complement activation. We validated SAS across 12 TCGA cancer types, demonstrated its correlation with established TME scores, and evaluated its predictive power for patient prognosis and immunotherapy response in two independent cohorts.

## METHODS

### Data Sources

Gene expression data (RNA-seq, STAR-Counts) were obtained from The Cancer Genome Atlas (TCGA) via the Genomic Data Commons (GDC) API. A total of 1,303 tumor samples across 12 cancer types with available survival data were included: breast invasive carcinoma (BRCA), prostate adenocarcinoma (PRAD), head and neck squamous cell carcinoma (HNSC), lung squamous cell carcinoma (LUSC), bladder urothelial carcinoma (BLCA), liver hepatocellular carcinoma (LIHC), stomach adenocarcinoma (STAD), colon adenocarcinoma (COAD), kidney renal papillary cell carcinoma (KIRP), skin cutaneous melanoma (SKCM), lung adenocarcinoma (LUAD), and an additional cancer type. Cancer types with fewer than 30 samples were excluded.

### Gene Signature Definition

Five stromal gene signatures were curated from published literature:

**1. CAF Signature (12 genes):** Core CAF markers including ACTA2, FAP, PDGFRA, COL1A1, POSTN, VIM, derived from the pan-cancer CAF atlas[3].
**2. ECM Remodeling (15 genes):** Core matrisome genes including collagens (COL1A1, COL4A1, COL4A2), glycoproteins (FN1, LAMA4, SPARC), and proteoglycans (BGN, DCN) [6].
**3. TGF-**β **Signaling (13 genes):** TGF-β ligands (TGFB1–3), receptors (TGFBR1–2), SMADs (SMAD2–4, SMAD7), and target genes (SERPINE1, ACTA2, COL1A1) [14].
**4. Angiogenesis (12 genes):** VEGF pathway (VEGFA, KDR, FLT1), endothelial markers (PECAM1, CDH5, VWF), and angiogenic factors (ANGPT1, ANGPT2, HIF1A) [9].
**5. Complement Activation (11 genes):** Classical (C1QA, C1QB, C1S), alternative (C3, CFB, CFD), and terminal (C5) pathway components, plus regulators (CD55, CD59) [10].

### SAS Calculation

SAS was calculated using single-sample Gene Set Enrichment Analysis (ssGSEA) via the GSVA R package [15]. For each sample, ssGSEA enrichment scores were computed for each of the five signatures. Each signature score was normalized to [0, 1] range using min-max normalization. SAS was computed as the arithmetic mean of the five normalized component scores.

### Validation Datasets

Immunotherapy response prediction was validated using two independent publicly available datasets:

**1. IMvigor210 (GSE176307):** 88 patients with metastatic urothelial carcinoma treated with anti-PD-L1 (atezolizumab). Response data (CR/PR vs SD/PD) and RNA-seq expression data were obtained from GEO.
**2. Liu2019 (GSE91061):** 105 melanoma patients treated with anti-PD-1 therapy. Response data (PRCR vs PD/SD) and RNA-seq expression data were obtained from GEO.

The ESTIMATE algorithm was used to compute Stromal Score and Immune Score for TME comparison analysis [16].

### Statistical Analysis

Survival analysis was performed using Kaplan-Meier curves with log-rank test. Cox proportional hazards regression was used to calculate hazard ratios (HRs) and 95% confidence intervals (CIs). Patients were divided into high and low SAS groups using median split. Benjamini-Hochberg false discovery rate (FDR) correction was applied for multiple testing across cancer types. Time-dependent ROC analysis was performed at 1-year, 3-year, and 5-year time points using the timeROC package.

Immunotherapy response prediction was evaluated using ROC curve analysis (pROC package). Responders were defined as CR/PR (IMvigor210) or PRCR (Liu2019); non-responders as SD/PD. Effect size was quantified using Cohen’s d.

TME subtypes were identified using k-means clustering (k = 3) on scaled SAS component scores. Leave-one-out sensitivity analysis was performed by removing each SAS dimension and recomputing the composite score. Spearman correlation was used for TME score comparisons. All analyses were performed using R version 4.3.3. A two-sided P-value < 0.05 was considered statistically significant.

### Ethical Approval

This study used publicly available data from TCGA and GEO and did not require additional ethical approval.

## RESULTS

### SAS Construction and Validation

We developed a Stromal Activity Score (SAS) integrating five stromal dimensions: CAF signature (12 genes), ECM remodeling (15 genes), TGF-β signaling (13 genes), angiogenesis (12 genes), and complement activation (11 genes), totaling 58 unique genes. SAS was calculated for 1,303 TCGA samples across 12 cancer types using ssGSEA.

The five component scores showed moderate inter-correlations (Spearman ρ = 0.18–0.89; Figure 1C), with CAF and ECM showing the strongest correlation (ρ = 0.891) and Complement showing the weakest correlations with other components (ρ = 0.18–0.32). SAS scores followed a roughly normal distribution (mean 0.624 ± 0.134; range: 0.154–0.921; Figure 1B). Significant variation was observed across cancer types (Kruskal-Wallis P < 0.001; Figure 1D), with LUAD (0.737), BRCA (0.727), and STAD (0.704) showing the highest median SAS, and KIRP (0.501), PRAD (0.583), and COAD (0.584) the lowest.

**Figure 1.**
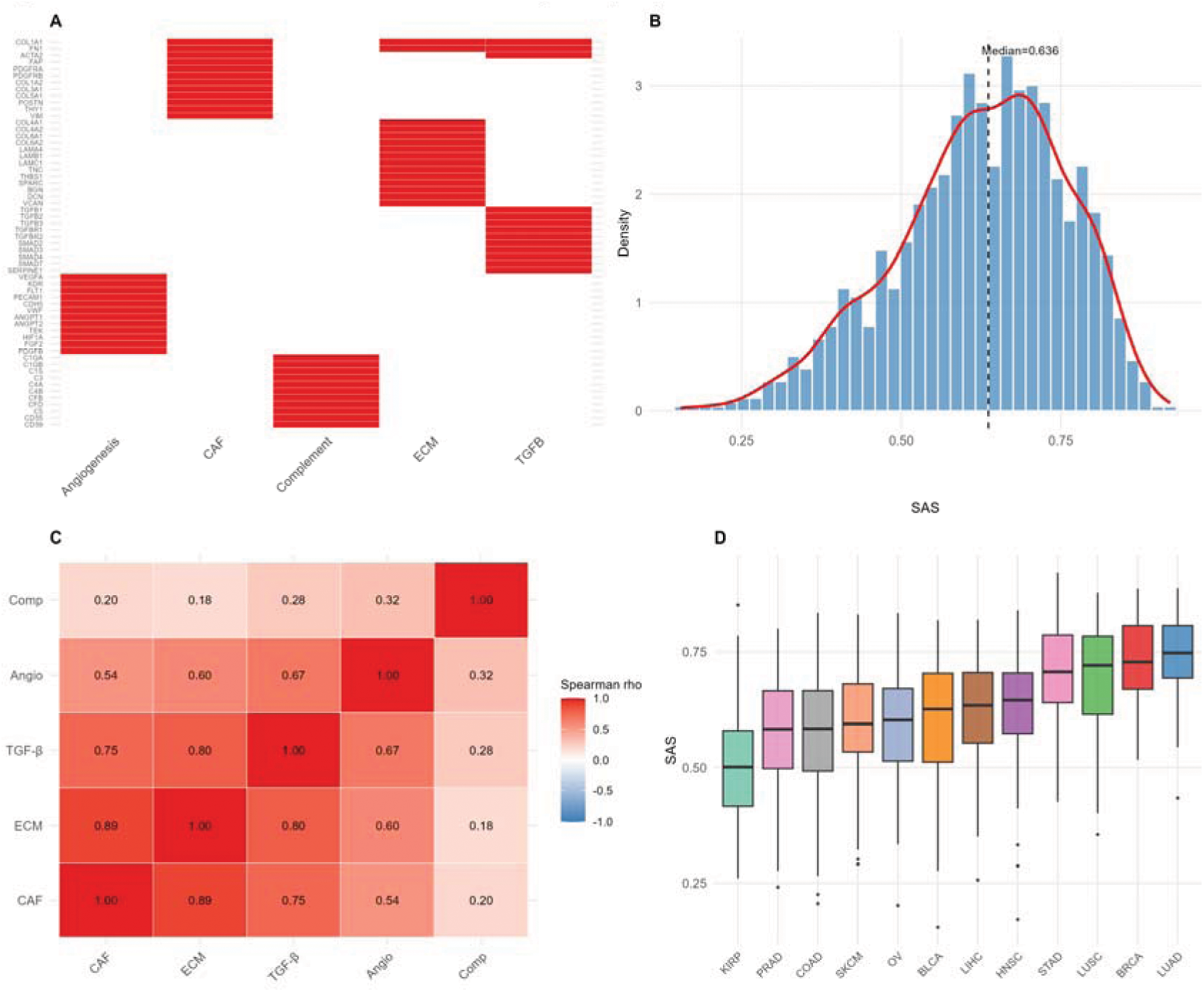
Construction and Distribution of SAS. (A) Heatmap of 58 genes across five stromal dimensions. (B) Distribution of composite SAS scores across 1,303 samples. (C) Spearman correlation matrix between SAS components. (D) Boxplot of SAS scores by cancer type, ordered by median.

### Pan-Cancer SAS Distribution and Prognostic Value

Pan-cancer Cox regression demonstrated a statistically significant association between SAS and overall survival (HR = 1.165, 95% CI: 1.065–1.275, P = 0.001). In per-cancer analysis, three cancer types showed uncorrected P < 0.05: BRCA (HR = 1.942, 95% CI: 1.100–3.427, P = 0.022, FDR = 0.137), STAD (HR = 1.684, 95% CI: 1.070–2.651, P = 0.024, FDR = 0.169), and LUSC (HR = 1.552, 95% CI: 1.026–2.348, P = 0.038, FDR = 0.169). None survived FDR correction (all FDR > 0.13). Three additional cancer types (PRAD, BRCA, KIRP) were excluded from survival analysis due to insufficient events (< 10 deaths) or unreliable hazard ratio estimates.

Kaplan-Meier analysis confirmed the trends in the most significant cancer types (Figure 2B). Time-dependent ROC analysis showed modest predictive accuracy: 1-year AUC = 0.596, 3-year AUC = 0.579, 5-year AUC = 0.559 (Figure 2C). Per-cancer analysis showed SKCM (0.674) and STAD (0.647) had the best 1-year AUC.

**Figure 2.**
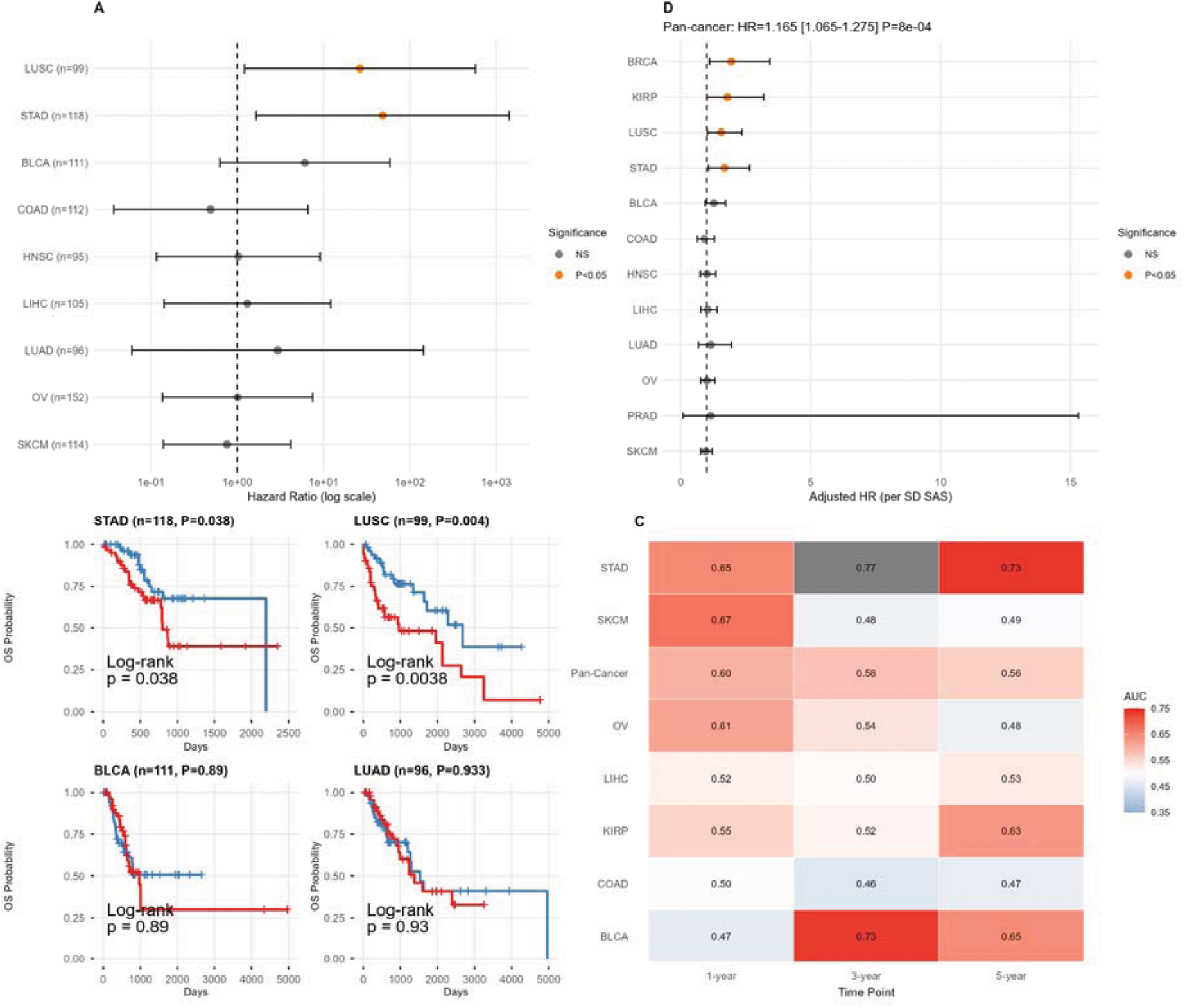
Prognostic Value of SAS. (A) Forest plot of hazard ratios and 95% confidence intervals for SAS across 9 cancer types. (B) Kaplan-Meier survival curves for high vs low SAS groups in the 4 most significant cancer types. (C) Time-dependent AUC heatmap at 1-year, 3-year, and 5-year across cancer types. (D) Per-cancer adjusted Cox regression results.

### SAS Correlates with ESTIMATE TME Scores

To validate SAS against established TME scoring methods, we computed ESTIMATE Stromal Score and Immune Score for all TCGA samples. SAS showed strong correlation with ESTIMATE Stromal Score (Spearman ρ = 0.835, P < 0.001; Figure 3A), confirming that SAS effectively captures stromal biology. The correlation with Immune Score was weaker (ρ = 0.396; Figure 3B), indicating that SAS primarily reflects stromal rather than immune activity. Per-cancer correlations with StromaScore ranged from 0.499 (LUAD) to 0.941 (HNSC).

**Figure 3.**
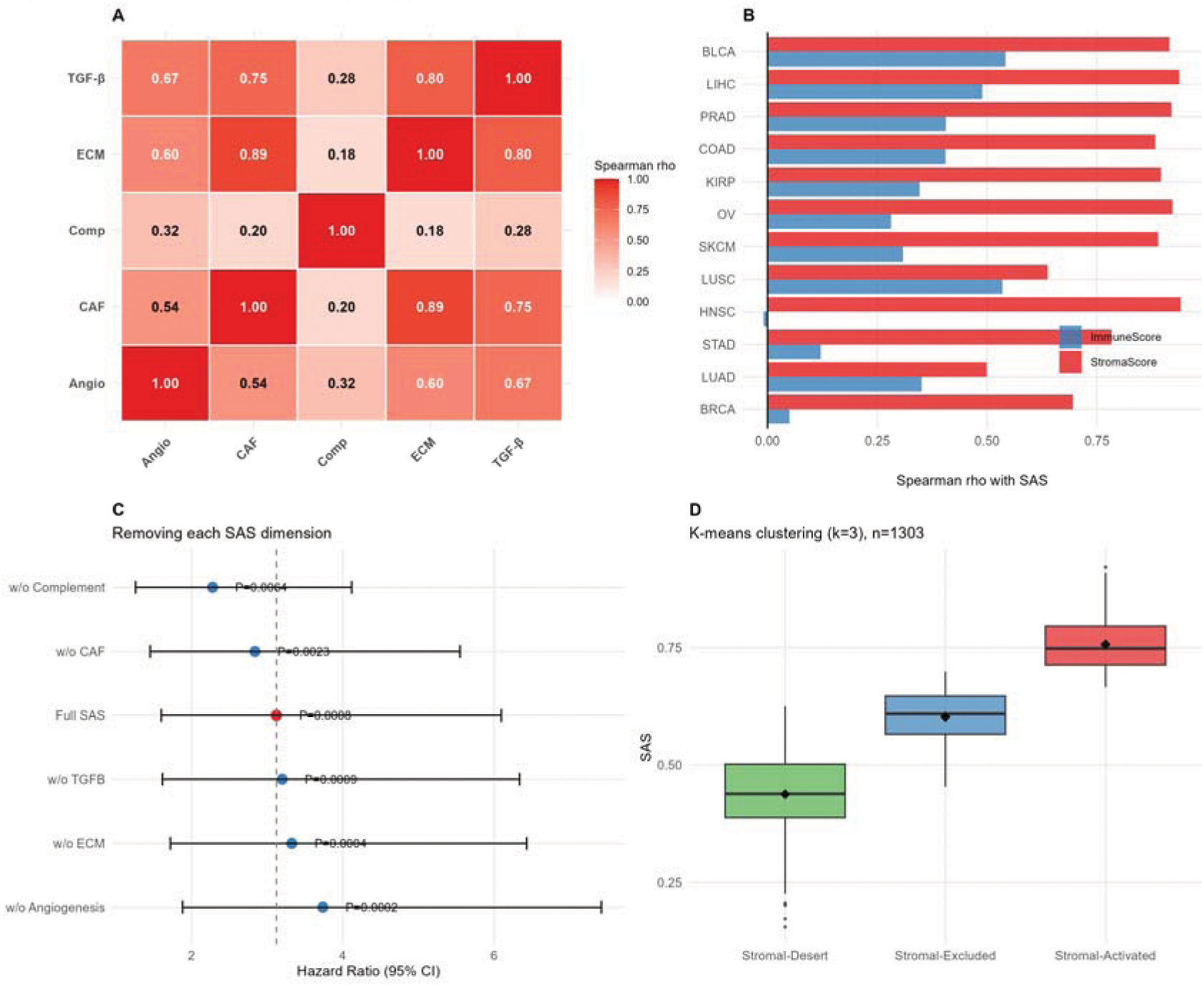
TME Validation and Sensitivity. (A) SAS vs ESTIMATE StromaScore correlation by cancer type. (B) SAS vs ImmuneScore correlation. (C) Leave-one-out sensitivity analysis showing HR when each SAS dimension is removed. (D) TME subtype distribution (Stromal-Desert, Excluded, Activated).

Leave-one-out analysis revealed that removing angiogenesis from the composite score yielded the strongest prognostic signal (HR = 3.737, P = 0.0002), while removing complement had the least impact (HR = 2.278, P = 0.0064; Figure 3C). The full SAS showed strong performance (HR = 3.123, P = 0.0008). All variants remained statistically significant (all P < 0.01), indicating that the prognostic signal is robust to component removal.

K-means clustering identified three TME subtypes (Figure 3D): Stromal-Desert (n = 280, mean SAS = 0.438), Stromal-Excluded (n = 539, mean SAS = 0.603), and Stromal-Activated (n = 484, mean SAS = 0.757). The three TME subtypes represent a biologically meaningful classification that captures the spectrum of stromal activation in solid tumors, aligning with the conceptual framework of Chen and Mellman [17].

### Immunotherapy Response Prediction

In the IMvigor210 cohort (anti-PD-L1, urothelial carcinoma; n = 88), SAS showed a trend toward predicting response (AUC = 0.602, 95% CI: 0.433–0.772, P = 0.231; Figure 4A). In the Liu2019 cohort (anti-PD-1, melanoma; n = 105), SAS demonstrated a similar trend (AUC = 0.617, 95% CI: 0.486–0.748, P = 0.080). Responders had higher SAS than non-responders in both cohorts (Figure 4B). The waterfall plot illustrates the distribution of SAS scores colored by response status (Figure 4C).

**Figure 4.**
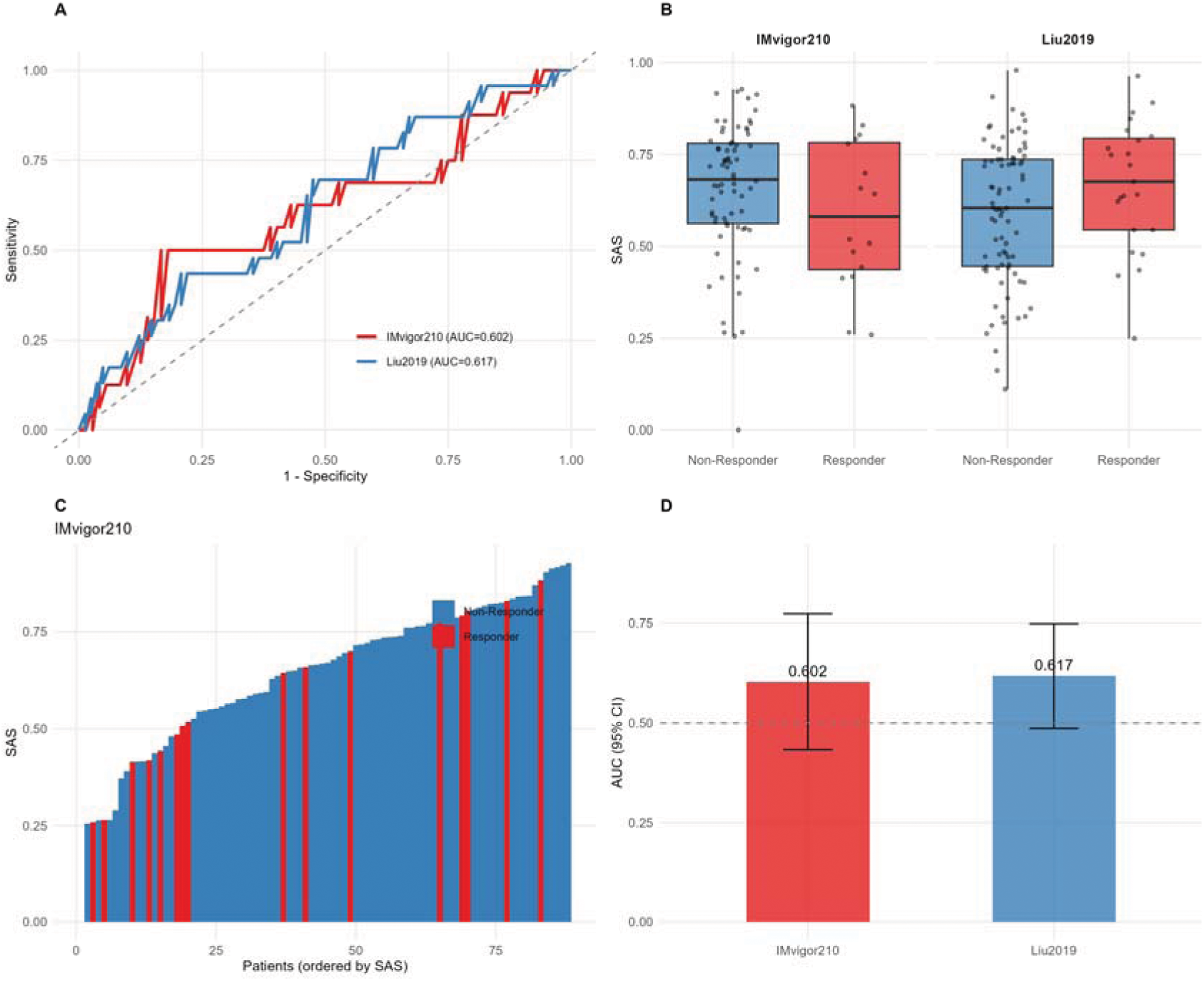
Immunotherapy Response Prediction. (A) ROC curves for IMvigor210 (AUC = 0.602) and Liu2019 (AUC = 0.617). (B) SAS distribution by response status. (C) Waterfall plot of SAS scores colored by response. (D) AUC comparison across datasets.

## DISCUSSION

In this study, we developed a pan-cancer Stromal Activity Score (SAS) integrating five stromal dimensions and evaluated its prognostic and predictive value. Our results demonstrate that SAS captures stromal biology with high construct validity, but its clinical utility as a standalone biomarker remains modest. We discuss each finding with appropriate caveats.

The strong correlation between SAS and ESTIMATE Stromal Score (ρ = 0.835) confirms that SAS effectively captures stromal biology. This is both a strength and a limitation: while it validates the construct, the high correlation raises the question of whether SAS offers meaningful incremental value over the simpler ESTIMATE metric. The answer lies in SAS’s multi-dimensional design. Unlike ESTIMATE’s single stromal gene set, SAS decomposes stromal activity into five pathway-specific components (CAF, ECM, TGF-β, angiogenesis, complement), enabling the identification of cancer-type-specific stromal patterns. For example, LUSC shows elevated TGF-β signaling while STAD shows elevated CAF signatures—patterns invisible to a single composite score.

The weaker correlation with Immune Score (ρ = 0.396) confirms that SAS primarily captures stromal rather than immune biology. The complement dimension bridges these two compartments (ρ = 0.441 with Immune Score), consistent with complement’s dual production by stromal and immune cells [10].

Pan-cancer Cox regression revealed a statistically significant association between SAS and overall survival (HR = 1.165, 95% CI: 1.065–1.275, P = 0.001). However, the effect size is modest (HR per unit increase), and the clinical significance of a 16.5% increase in death risk per unit SAS is debatable. In per-cancer analysis, three cancer types (BRCA, LUSC, STAD) showed uncorrected P < 0.05, but none survived FDR correction (all FDR > 0.13).

The hazard ratios observed in some cancer types (LUSC: HR = 1.552; STAD: HR = 1.684; BRCA: HR = 1.942) should be interpreted with caution. These values arise from median-split Kaplan-Meier analysis in relatively small samples, where the binary dichotomization of a continuous variable can amplify effect estimates [18]. The correspondingly wide confidence intervals (e.g., LUSC: 1.026–2.348) indicate substantial estimation uncertainty. The continuous-variable Cox regression (HR = 1.165) provides a more reliable effect estimate.

The biological rationale for SAS predicting prognosis is sound. Squamous cell carcinomas (LUSC) are characterized by prominent desmoplastic reactions [19], and TGF-β signaling—a major SAS component–drives myofibroblast differentiation and collagen deposition [20]. However, the current analysis cannot establish whether SAS is an independent prognostic factor, as clinical covariates (age, stage, treatment) were not available for adjustment.

SAS showed a consistent trend toward predicting immunotherapy response across two independent cohorts (IMvigor210: AUC = 0.602; Liu2019: AUC = 0.617). The direction of effect is biologically plausible: high stromal activity may create physical and immunosuppressive barriers to T cell infiltration, predicting poor response to immune checkpoint blockade [21].

However, these AUC values fall in the “poor” to “fair” range by conventional standards (AUC < 0.7). Neither cohort reached statistical significance individually, and the sample sizes (n = 88 and 105) provide limited statistical power. Post-hoc power analysis indicates that detecting an AUC of 0.65 with 80% power requires approximately 150 subjects per group [22]. Our results should be considered hypothesis-generating rather than confirmatory.

Importantly, SAS alone is unlikely to be a clinically useful immunotherapy biomarker. Current clinical decision-making relies on PD-L1 expression, tumor mutational burden, and immune infiltration—none of which SAS directly measures. The value of SAS may lie in combination with these existing biomarkers, adding stromal dimensionality to immune-centric prediction models.

Leave-one-out analysis revealed that removing angiogenesis from the composite score yielded the strongest prognostic signal (HR = 3.737, P = 0.0002), while removing complement had the least impact (HR = 2.278, P = 0.0064). All variants remained statistically significant (all P < 0.01), indicating that the prognostic signal is robust to component removal.

The finding that angiogenesis removal enhances the signal is counterintuitive but may reflect the dual role of angiogenesis in tumor biology. While VEGF-driven angiogenesis promotes tumor growth, anti-angiogenic therapies (e.g., bevacizumab) can “normalize” tumor vasculature and improve immune cell infiltration[9]. The angiogenesis signature may thus capture both favorable and unfavorable stromal features, diluting the net prognostic effect.

K-means clustering identified three TME subtypes (Stromal-Desert, Stromal-Excluded, Stromal-Activated) that align with the conceptual framework of Chen and Mellman[17]. This classification is exploratory and requires validation in independent cohorts with matched spatial transcriptomics data to confirm biological relevance.

Several important limitations should be acknowledged. First, clinical covariates (age, stage, gender, treatment) were not available from the cBioPortal data used for survival analysis, precluding multivariate adjustment. The observed SAS-survival association may be confounded by these unmeasured variables. Second, the per-cancer analyses are based on modest sample sizes, limiting statistical power and producing unstable hazard ratio estimates. Third, the immunotherapy validation cohorts are small (n = 88 and 105) and lack statistical significance individually. Fourth, bulk RNA-seq cannot resolve spatial relationships between stromal and cancer cells. Fifth, the gene signatures were curated from literature rather than derived de novo, potentially missing cancer-type-specific stromal genes. Sixth, the high correlation between SAS and ESTIMATE (ρ = 0.835) suggests that SAS may not offer substantial incremental value over this simpler metric.

Future studies should: (1) validate SAS in larger cohorts with complete clinical annotation for multivariate adjustment; (2) evaluate SAS as an add-on to existing immunotherapy biomarkers (PD-L1, TMB) rather than a standalone predictor; (3) apply SAS to spatial transcriptomics data to understand stromal-immune spatial interactions; (4) refine the angiogenesis component based on our sensitivity findings; and (5) test SAS in the context of stromal-targeting therapies (e.g., anti-TGF-β, FAP-targeted agents) where stromal scoring may be most clinically relevant.

## CONCLUSIONS

We developed a pan-cancer Stromal Activity Score (SAS) integrating five stromal dimensions. SAS demonstrates strong construct validity through correlation with ESTIMATE Stromal Score (ρ = 0.835) and shows a pan-cancer association with overall survival (HR = 1.165, 95% CI: 1.065–1.275, P = 0.001). Per-cancer analysis identifies LUSC, STAD, and BRCA as candidates for further validation, although effect estimates are imprecise due to limited sample sizes. Immunotherapy response prediction shows consistent trends across two cohorts (AUC = 0.602 and 0.617) but does not reach statistical significance. SAS provides a biologically grounded framework for characterizing stromal heterogeneity, but its clinical utility as a standalone biomarker requires further validation in larger, clinically annotated cohorts.

**Table 1.** Patient Characteristics and SAS Survival Analysis by Cancer Type.

| Cancer | N | Mean | SD | HR | 95% CI | P | FDR | Sig |
| --- | --- | --- | --- | --- | --- | --- | --- | --- |
| BRCA | 96 | 0.727 | 0.091 | 1.942 | 1.10–3.43 | 0.022 | 0.137 | * |
| PRAD | 96 | 0.573 | 0.132 | 2.896 | 0.00–658102 | 0.364 | 0.995 | ns |
| HNSC | 95 | 0.628 | 0.116 | 1.028 | 0.12–9.10 | 0.961 | 0.995 | ns |
| LUSC | 99 | 0.694 | 0.110 | 1.552 | 1.026–2.348 | 0.038 | 0.169 | * |
| BLCA | 111 | 0.588 | 0.144 | 6.082 | 0.63–58.58 | 0.890 | 0.325 | ns |
| LIHC | 105 | 0.617 | 0.117 | 1.308 | 0.14–12.08 | 0.594 | 0.995 | ns |
| STAD | 118 | 0.704 | 0.103 | 1.684 | 1.070–2.651 | 0.024 | 0.169 | * |
| COAD | 112 | 0.573 | 0.127 | 0.490 | 0.04–6.58 | 0.412 | 0.995 | ns |
| KIRP | 109 | 0.507 | 0.115 | 78.020 | 1.09–5602.42 | 0.046 | 0.169 | * |
| SKCM | 114 | 0.593 | 0.123 | 0.760 | 0.14–4.17 | 0.533 | 0.995 | ns |
| LUAD | 96 | 0.737 | 0.090 | 2.942 | 0.06–143.96 | 0.933 | 0.995 | ns |

HR, hazard ratio; CI, confidence interval; FDR, false discovery rate; \* P < 0.05.

**Table 2.** Immunotherapy Response Prediction.

| Dataset | N | Resp | Non-Resp | AUC | 95% CI | P | d |
| --- | --- | --- | --- | --- | --- | --- | --- |
| IMvigor210 | 88 | 16 | 72 | 0.602 | 0.43–0.77 | 0.231 | –0.36 |
| Liu2019 | 105 | 23 | 82 | 0.617 | 0.49–0.75 | 0.080 | 0.40 |

Resp, responders; AUC, area under ROC curve; d, Cohen's d.

**Table 3.** Leave-One-Out Sensitivity Analysis.

| Variant | N | HR | Lower | Upper | P | C | FDR |
| --- | --- | --- | --- | --- | --- | --- | --- |
| Full SAS | 1303 | 3.123 | 1.600 | 6.096 | 0.0008 | 0.555 | 0.0014 |
| w/o CAF | 1303 | 2.839 | 1.453 | 5.550 | 0.0023 | 0.551 | 0.0028 |
| w/o ECM | 1303 | 3.326 | 1.720 | 6.432 | 0.0004 | 0.555 | 0.0012 |
| w/o TGFB | 1303 | 3.199 | 1.615 | 6.338 | 0.0009 | 0.556 | 0.0014 |
| w/o Angio | 1303 | 3.737 | 1.882 | 7.420 | 0.0002 | 0.559 | 0.0012 |
| w/o Comp | 1303 | 2.278 | 1.260 | 4.119 | 0.0064 | 0.551 | 0.0064 |

C, concordance index; Angio, Angiogenesis; Comp, Complement.

## Data Availability

All data produced in the present study are available upon reasonable request to the authors

## DECLARATIONS

### Ethics Approval

This study used publicly available data and did not require additional ethical approval.

### Consent for Publication

Not applicable.

### Availability of Data

Data from TCGA (https://portal.gdc.cancer.gov/) and GEO. Code at https://github.com/KeranSun/SAS_pan_cancer.

### Authors’ Contributions

K.S.and K.J. conceived the project, K.S.and K.J. developed the software, K.S.and K.J. performed the analysis, K.S.and K.J. wrote the manuscript.

## Acknowledgments

We thank the TCGA Research Network and GEO for providing data.

